# Multivariable Mendelian Randomization Analysis Reveals the Protective Effect of Restless Legs Syndrome against Lower Limb Venous Diseases and Pulmonary Embolism

**DOI:** 10.1101/2024.08.27.24312632

**Authors:** Ru-Yu Zhang, Cheng Wen, Ru-Lin Geng, Jie-Hua Su, Xiao-Huan Liu, Zhuo-Ying Lai, En-Xiang Tao, Fu-Jia Li

## Abstract

**Objectives:** Lower limb immobility is a significant risk factor for peripheral vascular diseases, such as venous thromboembolism (VTE). Restless legs syndrome (RLS) is characterized by an irresistible urge to move the lower limbs to alleviate discomfort. The impact of RLS on lower limb vascular diseases is unclear.

**Methods:** This study used a two-sample Mendelian Randomization (MR) approach to assess the effect of RLS on the risk of seven arterial and seven venous diseases. Conducted in three stages, the study used different RLS datasets for discovery (Didriksen et al.) and replication (EU-RLS-GENE consortium) analyses across all the stages to ensure robust conclusions. Univariable MR (UVMR) was applied to analyze the causal relationship between RLS and 14 vascular diseases. Multivariable MR (MVMR) included covariates like BMI, smoking status, alcohol intake, and educational attainment. In the final stage, accelerometer-based physical activity at 12 different times were incorporated in MVMR analysis to determine if RLS influences lower limb vascular diseases by increasing limb activity.

**Results:** RLS was associated with a reduced risk of six venous diseases, including varicose veins, superficial phlebitis, VTE, and pulmonary embolism. These findings remained robust after adjusting for covariates but lost significance after accounting for nighttime physical activity.

**Conclusion:** RLS may lower the risk of venous diseases and pulmonary embolism by increasing nocturnal limb activity. This finding is particularly relevant for pregnant women and individuals with renal impairment, who are at high risk for both RLS and VTE. Further research is needed to validate these conclusions.

## Introduction

Venous thromboembolism (VTE), encompassing deep venous thrombosis (DVT) and pulmonary embolism, ranks as the third most common cardiovascular disease, following acute myocardial infarction and stroke, with a persistently high incidence rate^1^. VTE significantly affects patient survival and quality of life. Previous studies have identified hypercoagulability, venous stasis, and vascular injury as primary etiological factors ^2^.

Consequently, VTE often occurs secondary to trauma, prolonged immobilization, pregnancy, renal impairment, and malignancy. Lower limb immobility is a major risk factor for VTE, likely due to reduced muscle pump function and decreased blood flow velocity.

Restless Legs Syndrome (RLS) is a common sensorimotor disorder affecting 3.9% to 15% of the general population^3^. It is characterized by an overwhelming urge to move the limbs, typically manifesting in the evening or at night. Over 80% of RLS patients experience periodic leg movements in sleep (PLMS) ^4^. The frequency and severity of RLS symptoms vary widely; most patients experience mild symptoms, with only 11.9% seeking medical advice and approximately 3.4% requiring pharmacological treatment^5^. While previous research has primarily focused on the impact of RLS on sleep patterns, mood, daytime functioning, and cardiovascular health, the potential influence of increased nocturnal limb activity on VTE risk remains underexplored.

Pregnant women and patients with renal impairment are simultaneously exposed to the risks of VTE and RLS. About one-fifth of pregnant women develop RLS due to increased iron demands and hormonal changes, a prevalence rate two to three times higher than in the general population^6^. Concurrently, the risk of VTE in pregnant women is five times higher than in the general population due to hypercoagulability, hormonal fluctuations, and mechanical compression by the uterus^7^. For chronic kidney disease patients, particularly those on dialysis, the prevalence of RLS is approximately 20% to 30% due to iron and dopamine metabolism disorders. Additionally, renal impairment significantly elevates VTE risk, with incidence inversely related to kidney function ^8,9^. Notably, these patient groups face increased risks and challenges in the pharmacological treatment of both RLS ^10,11^ and VTE ^7,12,13^. Understanding the relationship between RLS and VTE is crucial for developing better treatment strategies for patients with comorbid risks, and for a more comprehensive understanding of the health outcomes associated with RLS.

Mendelian Randomization (MR) is a statistical method that uses single nucleotide polymorphisms (SNPs) as instrumental variables to infer causality ^14^. Compared to observational studies, MR results are less prone to confounding and reverse causality ^15^. Unlike randomized controlled trials (RCTs), MR studies do not require too stringent inclusion criteria, making their findings more reflective of real-world conditions ^16^. In this study, we employed MR to investigate the impact of RLS on lower limb vascular diseases.

This study comprises three stages. In the first stage, we conducted bidirectional univariable Mendelian randomization (UVMR) analyses to explore the causal relationships between RLS as the exposure and seven arterial and capillary diseases, along with seven venous system diseases, including VTE, as outcomes. In the second stage, we applied multivariable Mendelian randomization (MVMR) to adjust for confounding variables related to body mass index (BMI), smoking status, alcohol intake frequency, and years of education, thereby testing the robustness of our findings. In the third stage, we used MVMR again to examine the mediating role of limb activity at different times in the relationship between RLS and lower limb vascular diseases. Notably, we employed two independent datasets for discovery and replication analyses in each stage to ensure the robustness of our conclusions.

## Method

### 1. Study Design

The study design, as illustrated in **Figure 1**, consists of three stages. In the first stage, we employed UVMR analysis to investigate the impact of RLS on 14 lower limb vascular diseases. MR analysis requires three core assumptions: (1) the genetic instrumental variable (IV) must be strongly associated with the exposure; (2) the genetic IV must not be associated with confounders; (3) the genetic IV should influence the outcome only through the exposure^17^. Therefore, in the second stage, to minimize potential pleiotropy arising from BMI, smoking status, alcohol consumption frequency, and years of schooling, we included these traits as covariates alongside RLS in a MVMR analysis to further assess the robustness of our findings. In the third stage, we further used the MVMR approach to adjust for accelerometer-based physical activity at different times of the day to explore whether RLS indirectly influences the occurrence of lower limb vascular diseases by increasing nocturnal limb activity. Notably, in each stage, we conducted both discovery analysis (using RLS data from Didriksen et al. ^18^) and replication analysis (using RLS data from the EU-RLS-GENE consortium) to test the robustness of our conclusions. The study adheres to the STROBE-MR guidelines^19^ (**Table S2**).

**Figure 1.**
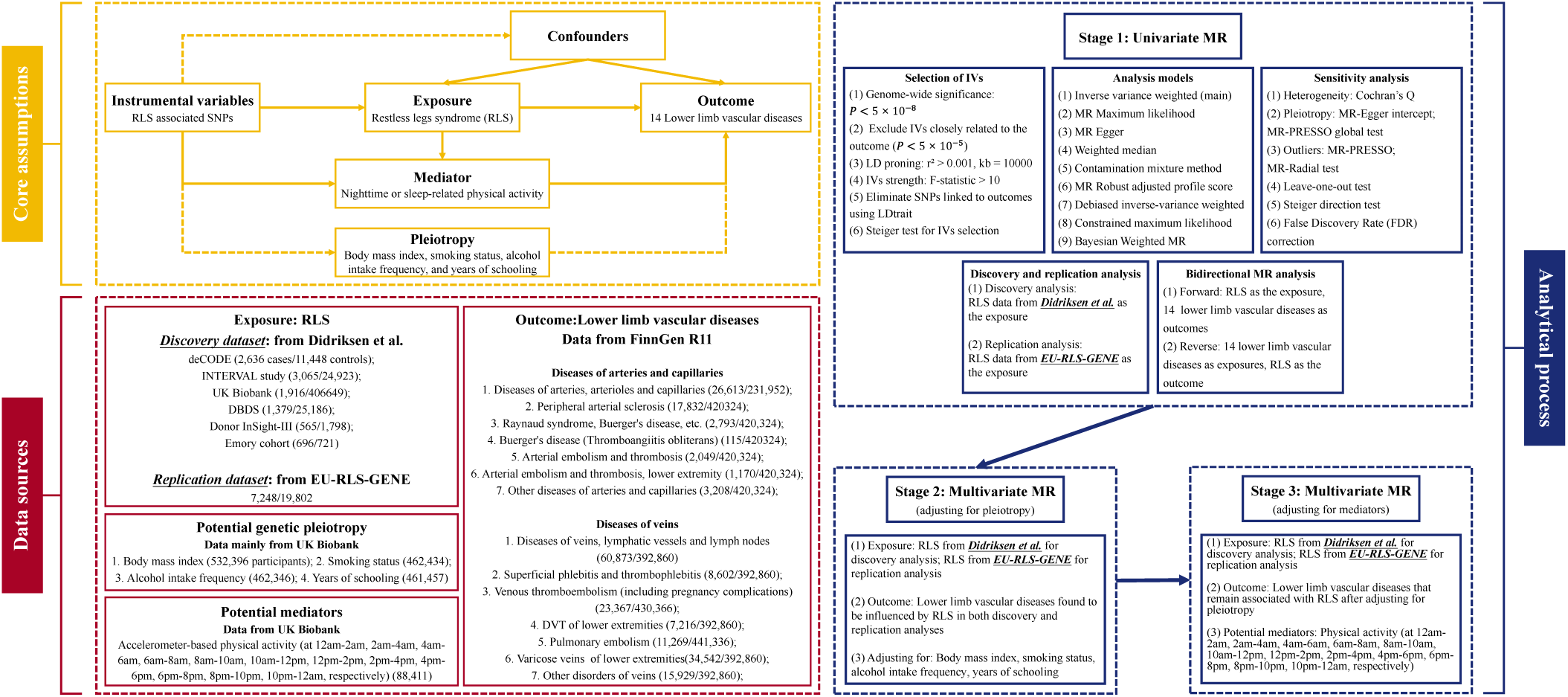
The study design. DVT, deep venous thrombosis. MR, Mendelian Randomization.

### 2. Data Sources

All data used in this study were derived from European populations. To enhance statistical power and the robustness of our conclusions, we selected the largest and most recent publicly available GWAS data for all traits. To minimize bias from sample overlap, the datasets for exposure and outcome factors were sourced separately.

#### 2.1 RLS

Genome-wide association study (GWAS) data for the discovery analysis were obtained from six independent cohorts, comprising a total of 480,982 participants (10,257 cases and 470,725 controls) ^18^. These include: (1) the deCODE cohort (2,636 cases and 11,448 controls), (2) the INTERVAL study (3,065 cases and 24,923 controls) (3) the DBDS cohort (1,379 cases and 25,186 controls), (4) the UK Biobank cohort (1,916 cases and 406,649 controls), (5) the Donor InSight-III cohort (565 cases and 1,798 controls), and (6) the Emory cohort (696 cases and 721 controls). RLS diagnosis in these studies was primarily based on the International RLS Study Group (IRLSSG) diagnostic criteria and derivative questionnaires, or was clinically diagnosed, supplemented by objective PLMS measurements (**Table S1**). Quality control, genotype imputation, and association testing were conducted at the cohort level, adjusting for age, sex, and principal components, followed by meta-analysis. The validation dataset was primarily derived from the international EU-RLS-GENE consortium (7,248 cases and 19,802 controls). RLS cases were diagnosed by neurologists or sleep specialists based on IRLSSG diagnostic criteria through face-to-face interviews (**Table S1**). Age, sex, and the top ten principal components from MDS analysis were included as covariates in the analysis.

### 2.2 Lower Limb Vascular Disease

To minimize interference from sample overlap, since some RLS data were sourced from the UK Biobank, we primarily used the latest and largest sample of lower limb vascular disease GWAS data from the FinnGen R11 database (https://r11.finngen.fi/)^20^. The FinnGen study combines genetic data from nationwide biobanks with disease status information from structured national healthcare databases. These data were adjusted for sex, age, genotype batch, and ten principal components. This study included seven arterial and capillary diseases (“diseases of arteries, arterioles, and capillaries”; “peripheral arterial atherosclerosis”; “Raynaud syndrome, Buerger’s disease, etc.”; “Buerger’s disease”; “arterial embolism and thrombosis”; “arterial embolism and thrombosis, lower extremity”; and “other diseases of arteries and capillaries”) and seven venous diseases (“diseases of veins, lymphatic vessels and lymph nodes”; “superficial phlebitis and thrombophlebitis”; “venous thromboembolism [including pregnancy complications]”; “DVT of lower extremities”; “pulmonary embolism”; “varicose veins of lower extremities”; and “other disorders of veins”) as outcomes. Sample sizes and diagnostic codes for these 14 vascular disease traits are detailed in **Table S1**.

### 2.3 Covariate and mediator

Data for BMI (532,396 individuals), smoking status (462,434 individuals), alcohol intake frequency (462,346 individuals), and years of schooling (461,457 individuals) were obtained from the most recent publicly available data from the UK Biobank. Data for physical activity at different time were also sourced from the UK Biobank. Previous studies have extensively used self-reported physical activity data, which may not accurately capture the timing of activity and can be influenced by cognitive and emotional factors ^21^. To objectively assess physical activity, UK Biobank invited participants to wear an accelerometer for 7 days, continuously recording physical activity data. The data were further quality controlled by Guanghao Qi and his team, ultimately including data from 88,411 participants ^22^. The relevant models adjusted for age, sex, and the top 20 genetic principal components. Individual physical activity levels were described over 12 non-overlapping two-hour periods throughout the day (measured by total acceleration logs for the periods: 12 am-2 am, 2 am-4 am, …, 10 pm-12 am).

## 3. Statistical Analysis

### 3.1 Stage one: Univariable MR Analysis

Genetic variants significantly associated with the exposure (P < 5 × 10^−8^) were selected as IVs. The R² value of SNPs estimated the variance in the exposure, and the F-statistic measured the strength of the instrumental variables. This study only included SNPs with an F-statistic greater than 10. Independent SNPs were identified through clumping based on European ancestry reference data (1000 Genomes Project, r² > 0.001, region = 10,000 kb). To satisfy the independence assumption of MR analysis, SNPs associated with the outcome were excluded based on LDtrait (**Table S4**, https://ldlink.nih.gov/?tab=ldtrait).

Inverse variance weighting (IVW)^23^ was our primary analysis method, as it offers precision and statistical power when all selected SNPs are valid IVs. To ensure robustness, we also applied eight secondary MR methods: Maximum Likelihood(ML) estimation^24^, Weighted Median method (WM)^25^, MR-Egger regression^26^, constrained Maximum Likelihood-Model Averaging (cML-MA) ^24^, Contamination Mixture method (ConMix) ^27^, Robust Adjusted Profile Score (MR-RAPS) ^28^, debiased IVW (dIVW), and Bayesian Weighted MR analysis (BWMR) ^29^.

To clarify the direction of causality, we implemented the following measures: (1) excluding IVs significantly associated with the outcome (P < 5 × 10^−5^)^30^, (2) further filtering IVs using the Steiger directionality test^31^, and (3) conducting reverse MR analysis.

To ensure the robustness of the results, we employed several stringent sensitivity analyses: (1) all three stages of analysis used two completely independent RLS datasets (discovery and replication datasets), and a result was considered credible only if both discovery and replication analyses were statistically significant; (2) to reduce false positives from multiple comparisons, we applied False Discovery Rate (FDR) correction to the p-values of individual analysis results; (3) robustness was ensured through the use of one primary and eight secondary MR methods; (4) heterogeneity in IVW estimates was assessed using Cochran’s Q statistic; (5) horizontal pleiotropy was evaluated using the MR-Egger intercept p-value^26^ and MR-PRESSO global test; (6) SNPs identified as outliers by MR-PRESSO or MR-radial were excluded, and the analysis was repeated with the remaining SNPs; (7) a leave-one-out analysis was performed to determine whether the results were driven by any single SNP.

MR results were expressed as odds ratios (ORs) with corresponding 95% confidence intervals (CIs). The estimates represent the change in ORs for lower limb vascular diseases per unit increase in the log value of RLS. In the reverse analysis, the results represent the relative OR increase in RLS per unit change in lower limb vascular diseases.

### 3.2 Stage 2 and Stage 3: MVMR Analysis adjusting for potential or mediator

To minimize potential pleiotropy due to BMI, smoking status, alcohol intake frequency, and years of schooling, we conducted MVMR analysis in stage 2 to evaluate the association between RLS and lower limb vascular disease risk after adjusting for these genetic traits. To determine whether RLS influences the incidence of lower limb vascular diseases through increased limb activity, we similarly conducted MVMR analysis in stage 3, estimating the causal effect of RLS on lower limb conditions after adjusting for physical activity during 12 periods throughout the day.

MVMR-IVW and MVMR-Egger methods were used to estimate causal effects. Pleiotropy was tested by comparing the MR-Egger intercept with zero. In the absence of pleiotropy, MVMR-IVW was the primary method for estimating MVMR effects; if pleiotropy was present, MVMR-Egger was used for estimation ^26^.

All analyses were two-sided and conducted using R software version 4.3.1, utilizing the TwoSampleMR^32^, MendelianRandomization^33^, MR-PRESSO^34^, RadialMR^35^, MR.RAPS^36^, MRcML^24^, BWMR^29^, and other R packages. All participating cohorts had obtained ethical approval, so no additional ethical approval or informed consent was required.

## Results

### 1. Stage 1: UVMR Estimation of RLS Impact on Lower Limb Vascular Diseases

In the discovery analysis, a total of 16 IVs were used, explaining 18% of the variance in RLS with F-values ranging from 1736.354 to 24514.887 (**Table S3**). In the replication analysis, 11 IVs were used, explaining 54% of the variance in RLS with F-values ranging from 248.334 to 10303.088 (**Table S3**). A search of the LD link database revealed no direct association between these IVs and the study outcomes (**Table S4**).

In the discovery analysis, after multiple comparison corrections, we identified seven significant causal relationships (**Figure 2A**). Genetically predicted RLS was associated with a decreased risk of “diseases of veins, lymphatic vessels, and lymph nodes” (OR [95% CI] = 0.941 [0.921, 0.962], P = 7.96E-08, P_fdr = 1.12E-06), “superficial phlebitis and thrombophlebitis” (OR [95% CI] = 0.867 [0.806, 0.932], P = 1.24E-04, P_fdr = 1.24E-03), “venous thromboembolism (including pregnancy complications)” (OR [95% CI] = 0.920 [0.890, 0.951], P = 8.30E-07, P_fdr = 5.81E-06), “DVT of lower extremities” (OR [95% CI] = 0.905 [0.844, 0.971], P = 5.21E-03, P_fdr = 1.74E-02), “pulmonary embolism” (OR [95% CI] = 0.921 [0.877, 0.967], P = 9.63E-04, P_fdr = 4.81E-03), “varicose veins of lower extremities” (OR [95% CI] = 0.937 [0.911, 0.962], P = 2.43E-06, P_fdr = 4.86E-05), and “other disorders of veins” (OR [95% CI] = 0.951 [0.915, 0.988], P = 9.52E-03, P_fdr = 1.90E-02).

**Figure 2.**
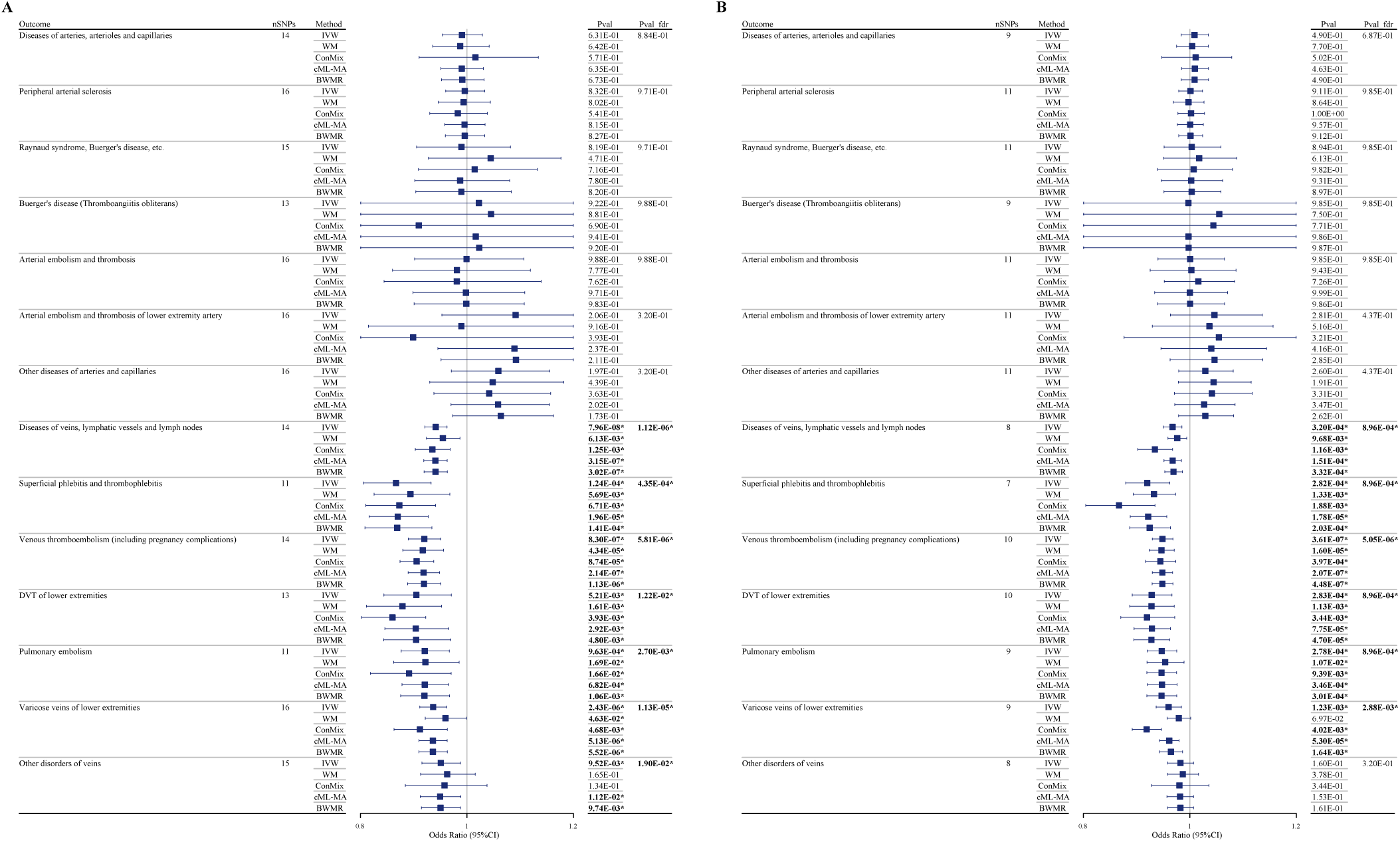
UVMR estimates of the impact of RLS on 14 lower limb vascular diseases. (A) Discovery analysis results. (B) Replication analysis results. nSNPs, number of instrumental variables; P_fdr, FDR-adjusted P-value. Statistically significant results are marked with an “*” and bolded.

Six of the seven causal relationships identified in the discovery analysis were successfully validated in the replication analysis (**Figure 2B**). Genetically predicted RLS was associated with a decreased risk of “diseases of veins, lymphatic vessels, and lymph nodes” (OR [95% CI] = 0.968 [0.950, 0.985], P = 3.20E-04, P_fdr = 8.96E-04), “superficial phlebitis and thrombophlebitis” (OR [95% CI] = 0.920 [0.880, 0.962], P = 2.82E-04, P_fdr = 8.96E-04), “venous thromboembolism (including pregnancy complications)” (OR [95% CI] = 0.949 [0.930, 0.968], P = 3.61E-07, P_fdr = 5.05E-06), “DVT of lower extremities” (OR [95% CI] = 0.928 [0.892, 0.966], P = 2.83E-04, P_fdr = 8.96E-04), “pulmonary embolism” (OR [95% CI] = 0.947 [0.920, 0.975], P = 2.78E-04, P_fdr = 8.96E-04), and “varicose veins of lower extremities” (OR [95% CI] = 0.960 [0.937, 0.984], P = 1.23E-03, P_fdr = 2.88E-03).

These findings were primarily based on IVW results. The remaining eight models showed results consistent with IVW in direction, with most reaching statistical significance (P < 0.05) (**Table S5**). All outliers were identified and excluded using MR-Radial and MR-PRESSO analyses. Cochran’s Q test indicated no heterogeneity in the results (**Table S6**). The MR-Egger intercept test and MR-PRESSO global test indicated that the results were not affected by horizontal pleiotropy (**Table S6**). The direction of the identified associations was confirmed by the MR Steiger test and reverse MR analysis (**Table S6, 8-10**). Leave-one-out analysis showed that these results were not driven by any single SNP (**Table S7**).

### 2. Stage 2: MVMR Adjusting for Genetic Pleiotropy

Based on the MVMR-IVW estimates, we found that when adjusting for genetically proxied BMI, smoking status, alcohol intake frequency, and years of schooling, RLS was still associated with a reduced risk of “diseases of veins, lymphatic vessels, and lymph nodes,” “superficial phlebitis and thrombophlebitis,” “venous thromboembolism (including pregnancy complications),” “DVT of lower extremities,” “pulmonary embolism,” and “varicose veins of lower extremities.” These results remained robust in both the discovery analysis (**Figure 3A**) and replication analysis (**Figure 3B**).

**Figure 3.**
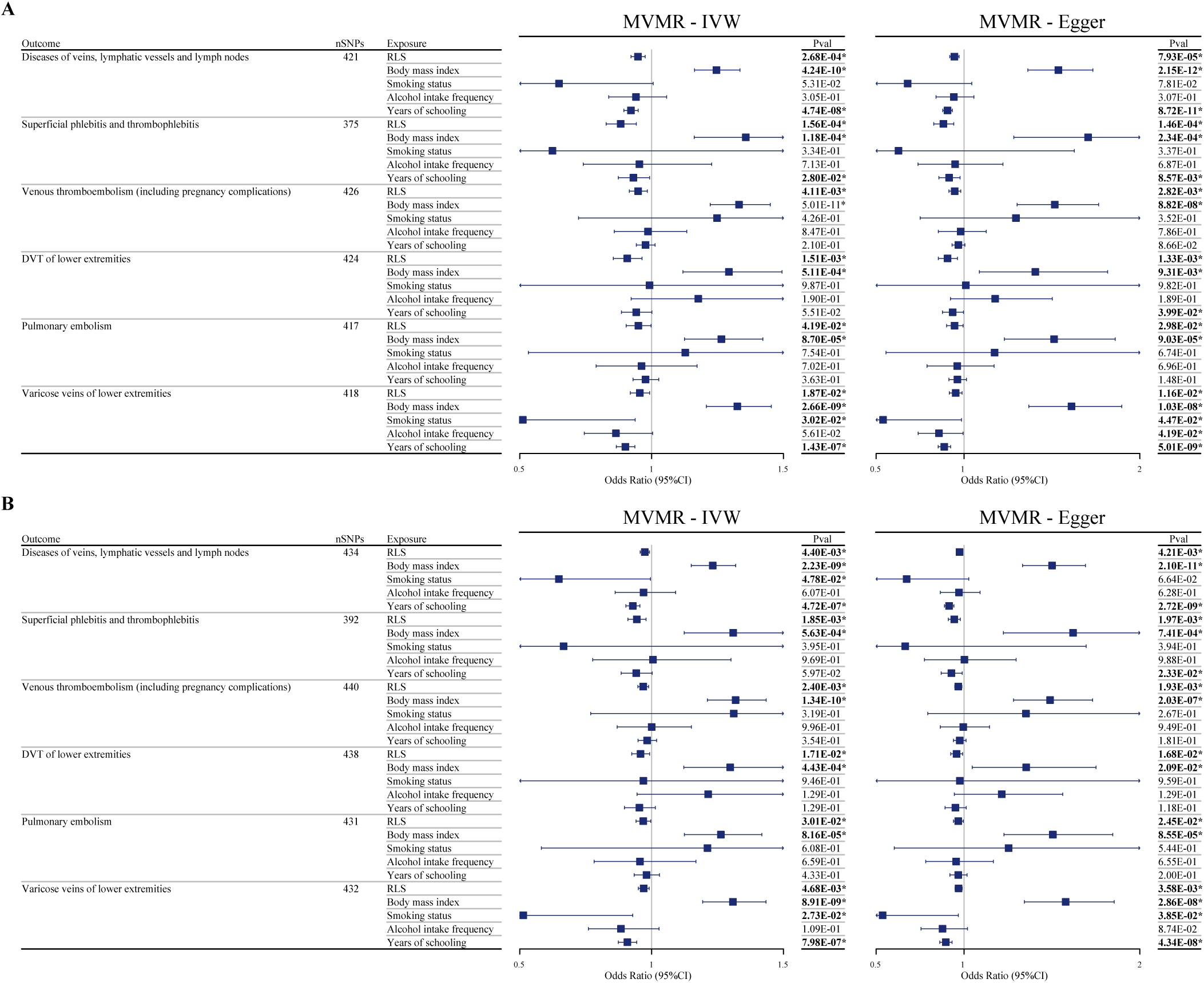
MVMR estimates of the impact of RLS on 6 lower limb vascular diseases after adjusting for covariates (estimated using MVMR-IVW and MVMR-Egger models). (A) Discovery analysis results. (B) Replication analysis results. RLS, Restless Legs Syndrome. Statistically significant results are marked with an “*” and bolded.

Sensitivity analysis suggested that some results might be influenced by heterogeneity and horizontal pleiotropy (**Table S11**). This could be due to the inclusion of too many covariates. In such cases, we referred to the MVMR-Egger results. Given that the direction of RLS effects was consistent and statistically significant across all analyses using both the MVMR-IVW and MVMR-Egger models, we consider the MVMR results to be robust and reliable.

### 3. Stage 3: MVMR Adjusting for Physical Activity at Different Time

In the discovery analysis, after adjusting for physical activity during the 12 am-2 am period, the effect of RLS on “diseases of veins, lymphatic vessels, and lymph nodes” was no longer significant. After adjusting for physical activity during the 12 am-2 am or 2 am-4 am periods, the effect of RLS on “venous thromboembolism (including pregnancy complications)” was no longer significant. After adjusting for physical activity during the 10 pm-12 am, 12 am-2 am, or 2 am-4 am periods, the effect of RLS on “DVT of lower extremities” was no longer significant. After adjusting for physical activity during the 8 pm-10 pm, 12 am-2 am, or 2 am-4 am periods, the effect of RLS on “pulmonary embolism” was no longer significant (**Figure 4**).

**Figure 4.**
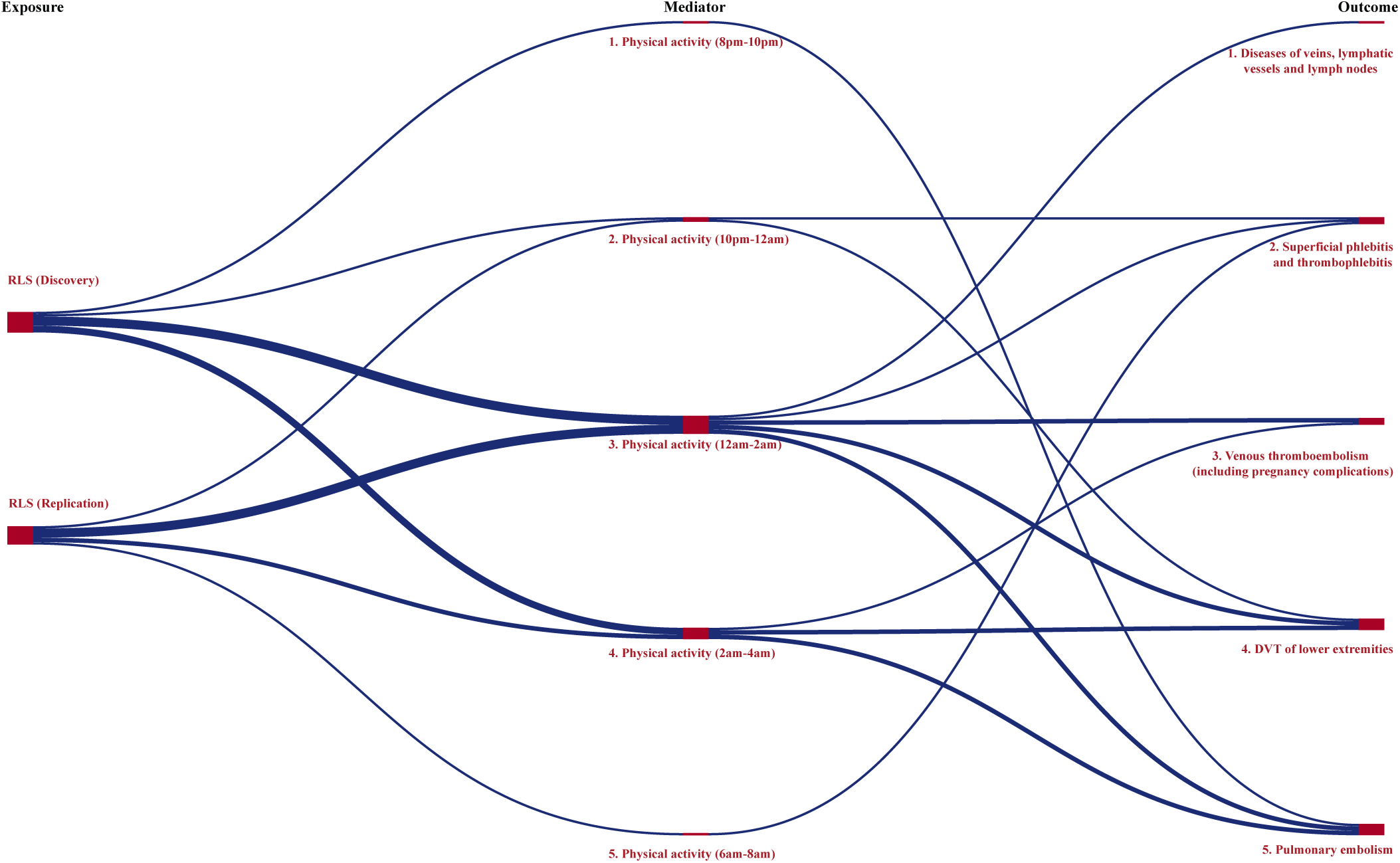
MVMR estimates of the mediating effect of physical activity at different time periods on the relationship between RLS and lower limb vascular diseases. If the impact of RLS on a given outcome is no longer significant after adjusting for physical activity during a specific time period, that period’s activity is considered a mediator. All mediating relationships are indicated by dark blue lines.

In the replication analysis, after adjusting for physical activity during the 10 pm-12 am, 12 am-2 am, or 6 am-8 am periods, the effect of RLS on “superficial phlebitis and thrombophlebitis” was no longer significant. After adjusting for physical activity during the 12 am-2 am period, the effect of RLS on “venous thromboembolism (including pregnancy complications)” was no longer significant. After adjusting for physical activity during the 12 am-2 am or 2 am-4 am periods, the effect of RLS on “DVT of lower extremities” was no longer significant. After adjusting for physical activity during the 12 am-2 am or 2 am-4 am periods, the effect of RLS on “pulmonary embolism” was no longer significant (**Figure 4**). For the MVMR results and sensitivity analyses of Stage 3, please refer to **Table S12** and **Table 13**.

## Discussion

This study systematically explored the impact of RLS on 14 peripheral vascular diseases across three stages. In the first stage, UVMR analyses revealed that genetically predicted RLS was associated with a reduced risk of the following conditions: “diseases of veins, lymphatic vessels and lymph nodes”; “superficial phlebitis and thrombophlebitis”; “venous thromboembolism [including pregnancy complications]”; “DVT of lower extremities”; “pulmonary embolism”; “varicose veins of lower extremities”. In the second stage, after adjusting for IVs related to BMI, smoking status, alcohol intake frequency, and years of education in the MVMR analysis, the results remained robust. In the third stage, after adjusting for physical activity during the 10 pm-12 am, 12 am-2 am, 2 am-4 am, and 6 pm-8 am periods, the effect of RLS on these outcomes was no longer significant. These findings were consistent and mutually supportive in both discovery and replication analyses.

RLS is a common sensorimotor disorder, affecting 3.9% to 15% of the general population^3^. Its primary symptoms include an urge to move the limbs that occurs or worsens at rest, typically in the evening or at night, with relief following movement. Over 80% of RLS patients experience PLMS. Symptom frequency and severity vary widely, with most patients experiencing mild symptoms; only 11.9% seek medical consultation, and approximately 3.4% require pharmacological treatment^5^. Previous research has mainly focused on the effects of RLS on sleep, quality of life, and mood. Some studies suggest that PLMS may lead to abnormal sympathetic activation, potentially increasing cardiovascular disease risk, although this conclusion remains controversial. Notably, increased limb activity might also promote blood circulation through the muscle pump mechanism, improving microcirculation. However, the impact of RLS on lower limb vascular diseases has not been thoroughly studied.

Through MR analysis, our study is the first to identify that genetically predicted RLS, by increasing limb activity during the evening/night (10 pm-12 am, 12 am-2 am, 2 am-4 am, or 6 pm-8 am), reduces the risk of six venous diseases (“diseases of veins, lymphatic vessels and lymph nodes”; “superficial phlebitis and thrombophlebitis”; “venous thromboembolism [including pregnancy complications]”; “DVT of lower extremities”; “pulmonary embolism”; “varicose veins of lower extremities”). Methodologically, compared to observational studies, MR has the advantage of being less susceptible to confounding and reverse causality. MR studies, unlike RCTs, are more reflective of real-world population distributions, making their conclusions more generalizable. From a results perspective, after FDR correction, the false positive rate was low, and the results were consistent across multiple computational models, unaffected by heterogeneity, pleiotropy, or outliers, and remained robust in MVMR adjustments. More importantly, the findings were validated in two completely independent datasets, indicating high reliability and warranting further investigation in future observational studies and clinical practice.

Venous stasis due to lower limb immobility is a key factor in venous thrombosis. While normal individuals have less limb activity during sleep, RLS patients exhibit increased limb movement, predominantly at night. Additionally, RLS symptoms commonly occur in specific situations such as during car rides, long flights, or prolonged sitting in cinemas or theaters. Although each patient has their own symptom relief strategies, the most common include walking, stretching, massaging the affected limb, or standing barefoot on the floor^37^. These behaviors can exert pressure on the lower limbs, enhancing the muscle pump effect, thereby promoting venous return, improving venous valve function, and ultimately reducing the risk of various lower limb venous diseases. The MVMR analysis indicated that after adjusting for nocturnal limb activity, the protective effect of RLS on venous diseases disappeared, further corroborating this point.

These findings may have significant implications for pregnant women and those with renal impairment, who are at risk for both RLS and VTE. Clinicians must carefully consider the risks of teratogenicity, bleeding, and renal function impairment when treating these patients^7,10,38^. If RLS indeed reduce the risk of VTE, could we manage RLS symptoms to a mild degree rather than completely eliminating them in patients who already have RLS and are at high risk for VTE, in order to harness its protective effect? After all, most non-pharmacological preventive measures involve daytime voluntary movement, while RLS symptoms primarily occur at night, potentially complementing each other in terms of activity timing. However, we must also exercise caution: severe RLS can impair sleep quality in pregnant women, potentially leading to mood disorders, prolonged labor, cesarean sections, or even preeclampsia^10,39,40^. Furthermore, severe RLS may increase the risk of muscle atrophy in end-stage renal disease patients and those on dialysis^41^, as well as cardiovascular events and mortality^42^. Therefore, more research is needed to clarify these issues.

The main strengths of this study lie in the robustness of findings. (1) Our study yielded consistent results across multiple datasets (exploratory and validation datasets), multiple models (one primary analytical method and eight secondary analytical methods), and multiple approaches (UVMR, MVMR, FDR correction, and other sensitivity analyses). (2) In the MVMR analysis, the data on limb activity periods were not subjectively reported by participants but were objectively recorded via wearable devices. However, we also acknowledge the following limitations: (1) All datasets used in the analysis were derived from European populations, which may limit the generalizability of the findings to other ethnic groups; (2) The study’s conclusions were based solely on MR analysis, lacking external validation from observational studies, preventing triangulation of evidence.

## Conclusion

Our study suggests that RLS, by increasing nocturnal limb activity, reduces the risk of lower limb venous diseases (especially VTE) and pulmonary embolism. This finding may have significant implications for pregnant women and those with renal impairment, who are at risk for both conditions. Further clinical research is needed to confirm these findings.

## Data Availability

All data produced in the present study are available upon reasonable request to the authors

## Acknowledgments

This study was supported by the National Natural Science Foundation of China (grant number 82271454) and the Basic Research Program of the Shenzhen Science and Technology Project (grant number JCYJ20220818102206014). We extend our sincere gratitude to all the participants for their valuable contributions.

---

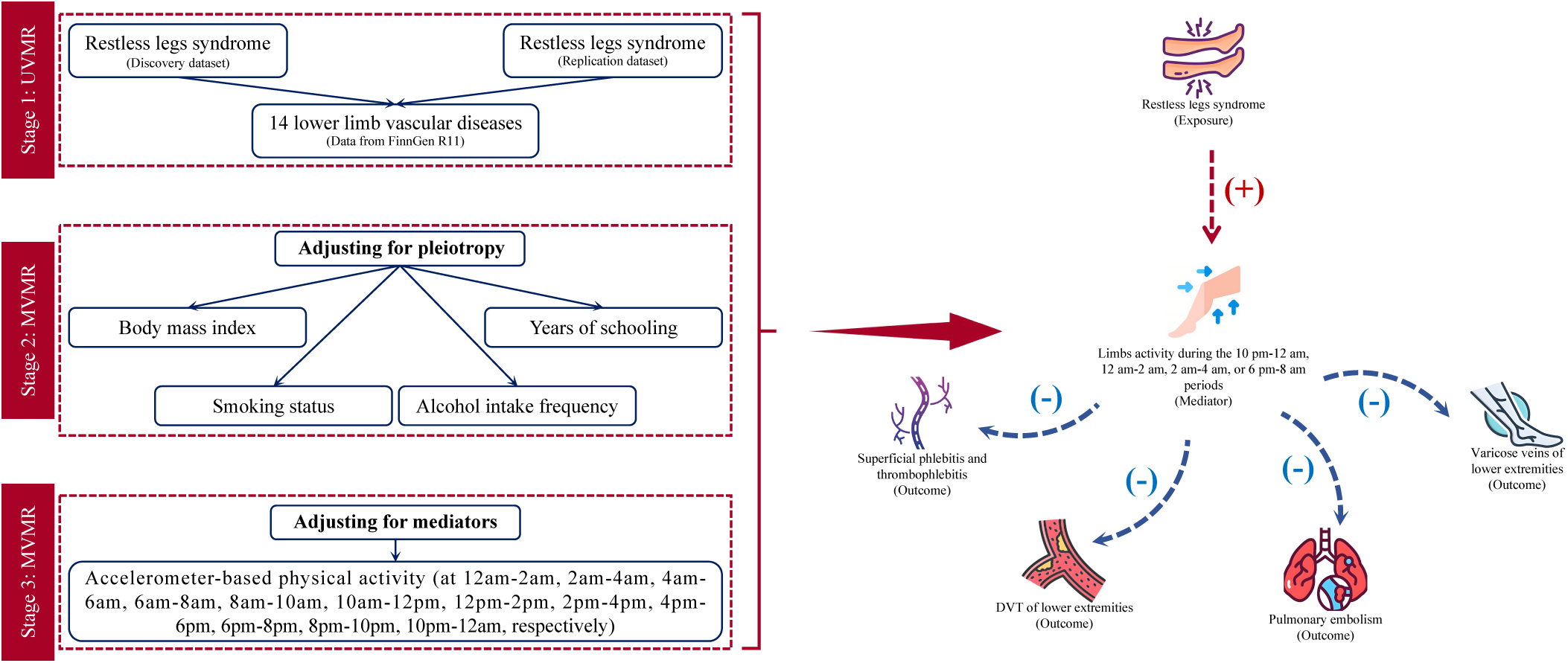

